# Mental Health and Time Management Behavior among Students During COVID-19 Pandemic: Towards Persuasive Technology Design

**DOI:** 10.1101/2021.10.01.21264409

**Authors:** Mona Alhasani, Ali Alkhawaji, Rita Orji

## Abstract

The study explored the impact of the COVID-19 pandemic on students’ mental health in higher education while capturing their perceptions and attitudes towards time management. The aim was to examine relationships between stress, anxiety, and specific time management related factors. Considering possible differences between genders and degree levels, we developed five structural equation models (SEMs) to delineate these relationships. Results of a large-scale study of 502 participants show that students suffered from stress and two types of COVID-19-related anxiety: disease and consequences. Students’ preference for organization was the only factor that significantly promoted their perceived control over time, which contributes to reducing stress, hence, anxiety. However, female students reported higher stress and anxiety levels than male students. Graduate students reported higher anxiety levels related to the consequences of the pandemic compared to undergrads. To promote students’ preference for organization, we map the three categories of organization to corresponding persuasive strategies which could be used in the design of persuasive interventions. This creates an opportunity for developing technological interventions to improve students’ perceived control over time, thus, reduce stress and anxiety.

## 1 INTRODUCTION

Since the declaration of the coronavirus (COVID-19) outbreak as a global pandemic, strict public health measures have been implemented, changing daily life in many countries around the world. During these times, the requirement of social distancing and self-quarantine has imposed unconventional lifestyles with which harmful habits may have developed, compromising the welfare of whole populations [42][5].

In higher education, the sudden transition from Face-to-Face to online teaching has introduced pedagogies that place students in complete charge of their learning. Moreover, the use of virtual platforms has widely increased to simulate classroom interaction in which synchronous learning is facilitated. Experiencing academic pressure in such educational environments can substantially contribute to students’ high stress levels, affecting their performance [7].

Stress is a major risk factor for serious illness. With adverse effects on various bodily organs, stress can impact student’s mental as well as physical and emotional well-being [15]. Since maintaining a balanced lifestyle is key to productivity and success, academic advising services often promote time management as an effective way to cope with stress [26].

### 1.1 Time management and its impact

Time management is one major issue among students in higher education; it influences how they perceive and utilize time in a way that allows them to juggle their academic tasks accordingly [26]. Students often complain about having heavy academic workloads that require more time than they have during the term. With congested daily schedules, students can feel overwhelmed, trying to meet all deadlines. Increased pressure of academic demands can create a stressful experience, especially as a result of disorganization [20].

Students waste time when they search for and fail to find important information. Spending time repeating tasks (such as checking e-mails, social media, etc.) and worrying about uncompleted tasks also waste time and impede progress. Therefore, students may find it difficult to distinguish between what is important and what is not. The ability to utilize time in an effective and efficient manner has been utilized as an effective approach to cope with stress [26].

Various time management strategies can be used to support effective time management Behavior [9]. Task prioritization strategy requires a distinction between important and urgent tasks [28]. In the Time Management Matrix, also known as Eisenhower’s Matrix (Figure 1), Covey et al. categorized tasks into four quadrants - urgent, not urgent, important, and not important - and suggested possible strategies for managing tasks depending on their urgency and importance [12]. The use of personal planning tools, such as electronic planners, pocket diaries, calendars, computer programs, wall charts, index cards, and notebooks, is another strategy recommended to improve work productivity [9]. Contrary to the common belief that multi-tasking saves time, studies in human psychology have shown the opposite is often the case. Swinging between tasks wastes time and impacts productivity [43]. When it becomes routine, multi-tasking may lead to difficulties to focus when needed [43].

**Fig. 1.**
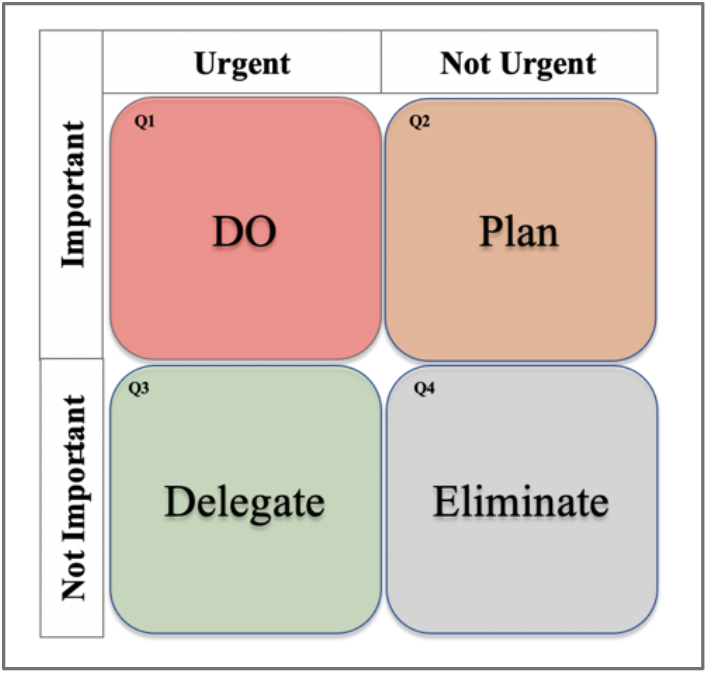
The Time Management Matrix (Eisenhower’s matrix)

Improper time allocation and last-minute cramming for exams are causes of stress and poor academic outcomes [31]. Practicing effective time management strategies, such as setting goals and priorities and monitoring progress over time [23], is expected to improve students’ perceived control over time [26]. Research shows that improved perceived control over time has a significant positive impact on students’ perceptions and attitudes toward work and life, promoting productivity and minimizing stress levels [23][26]. Exploring how perceived control over time influences students’ stress and various stress-related outcomes, Nonis et al. reported that students with high perceived control over time experienced lower academic stress levels and had higher academic performance and problem-solving abilities (i.e., approach-avoidance and personal control), which contributed to better health (i.e., physiological and psychological health) compared with those with low perceived control over time [32].

### 1.2 Persuasive Technology design

Persuasive technology (PT) are interactive systems that aim to aid users to achieve behavioral change by promoting and reinforcing desirable perceptions and attitudes [39]. Effective application of PT is growing in various areas of health and wellness [40][3][53]. The design of PT incorporates persuasive strategies (PSs), which are the backbone mechanics upon which PT operates to motivate desired behaviours.

Fogg provided seven strategies for designing a persuasive system [16], upon which Oinas-Kukkonen and Harjumaa built and proposed a framework for persuasive system design. The Persuasive Systems Design (PSD) framework comprises of 28 strategies for persuasive system design content and functionality [33]. Based on the type of support that they provide, these strategies were categorized into four categories: *primary task, dialogue, system credibility*, and *social support* [33]. (See Figure 2). Furthermore, Goal-Setting strategy has widely been employed in the design of persuasive systems as it has been proven to direct individuals’ attention and increase task performance [25][22]. This strategy is based on a theory that focuses on how individuals set up goals, how they react to them, and how they use them to achieve behavioural change [25].

**Fig. 2.**
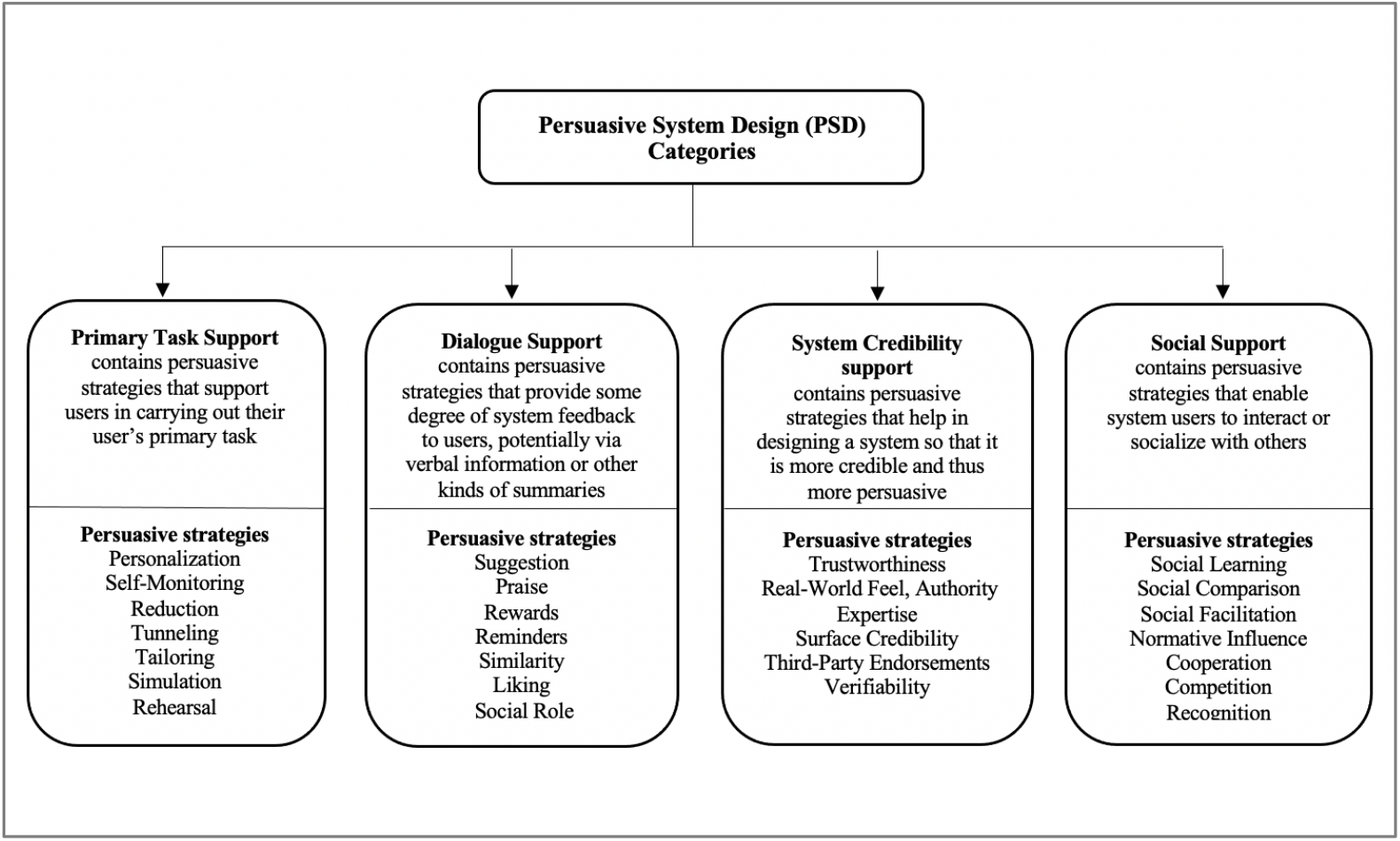
Framework for persuasive system design (PSD)

### 1.3 Aim of the study

Literature available on the impact of the COVID-19 pandemic on students’ mental health [24] [54] lacks insights into their respective time management behaviour. The study objective was to explore, from a global perspective, the impact of the COVID-19 pandemic on students’ mental health in higher education while capturing their perceptions and attitudes towards time management. The specific aim was to examine the relationships between stress, anxiety, and specific factors related to time management behavior, considering possible differences between genders and degree levels.

To address the study objective, we ask the following five questions:

**RQ 1:** Did students in higher education suffer stress and anxiety?

**RQ 2:** Are there any differences between students’ stress and anxiety levels based on their gender or degree level?

**RQ 3:** What are the most practiced time management behaviors among students?

**RQ 4:** Which time management factor is significantly associated with perceived control over time?

**Key findings** will provide insights that could guide persuasive technological interventions to support students in managing time and stress.

## 2 METHODOLOGY

In this section, we describe the study design, measurement instruments, and statistical analysis procedures.

### 2.1 Data Collection

Ethics approval was obtained from Research Ethics Board (REB) at Dalhousie University. The study was based on an online survey launched from 17 August 2020 to 30 January 2021. Students were recruited using different approaches via email (academic) and social media (Twitter, Facebook, and WhatsApp). Amazon Mechanical Turk (MTurk) was also used to recruit participants. MTurk is an accepted way to collect responses from diverse participants [17][18][38][35]. Research has shown that data obtained from MTurk are at least as reliable as those obtained via traditional methods even in clinical research [4][6][46]. Using the consent form, all participants were informed about the purpose of the study, data and privacy at the beginning of the survey.

### 2.2 Measurements

The measurement instruments used in this study will be delineated in this section.

#### 2.2.1 Perceived Stress Scale (PSS-10)

PSS-10 was developed by Cohen et al. [11]. The scale measures individuals’ perceived stress levels using ten 5-point Likert items. The items ranging from 0 (*never*) to 4 (*very often*) are intended to assess essential elements related to stress. The items specifically ask the respondents about their feelings and thoughts during the COVID-19 pandemic– how overloaded, uncontrollable, and unpredictable their lives were. The **total score** of the scale ranges from 0 to 40. Scores ranging from 0 to13 are considered as **low** self-perceived stress levels; scores from 14 to 26 are considered as **moderate** self-perceived stress levels and scores from 27– 40 are considered as **high** perceived stress levels. The scores of four items (4, 5, 7, and 8) were reversed according to the scale authors’ recommendations and usage guideline. An example of revered items is “how often have you felt confident about your ability to handle your personal problems” (item 4).

#### 2.2.2 Time Management Behavior (TMB) Scale

TMB scale was developed by Macan and Shahani (1990) to assess behaviors critical to time management [26]. The scale consists of 34 items, grouped into four categories. Each category represents one of four factors. Factor 1 - *Setting Goals and Priorities* (SGP, 10 items): refers to setting goals that individuals need to accomplish and prioritizing tasks to achieve the needed goals. Factor 2 - *Mechanics of Time Management* (MTM, 11 items): refers to managing time by planning, scheduling, and making lists. Factor 3 - *Preference for Organization* (PFO, 8 items): refers to the preference for an organization in an individual’s study space and approach to the assigned tasks. Factor 4 - *Perceived Control of Time* (PCT, 5 items): refers to the extent to which individuals believe they can directly affect how their time is spent.

The scale measures these time management factors using a 5-point Likert items ranging from 1 (seldom true) to 5 (very often true). The scores of negatively worded items were reversed. Higher **average** scores indicate more frequent use of time management behavior.

#### 2.2.3 Pandemic Anxiety Scale (PAS-9)

PAS-9 was developed by a panel of psychologists with expertise in mental health [29]. Using 5-point Likert item ranging from 1 (Strongly disagree) to 5 (Strongly agree), PAS-9 captures two types of anxiety related to COVID-19 pandemic using 9 items. The first 6 items capture anxiety driven by worries about the disease itself, such as “*I’m worried that I will catch COVID-19*”, while the last 3 items capture anxiety related to other consequences of the pandemic, such as “*I’m worried about the long-term impact [the pandemic] will have on my job prospects and the economy*.”

### 2.3 Statistical analysis

We summarize the steps taken to analyze our data, using various well-known analytical tools and procedures [36].

1. We calculated descriptive statistics using frequency, mean, and standard deviation.
2. We used Statistical Package for the Social Sciences (SPSS) version 23.0 to:
  a. Calculate the Kaiser-Meyer-Olkin (KMO) sampling adequacies and the Bartlett Test of Sphericity [19] to determine the suitability of our data for further analysis.
  b. Examine students’ stress, anxiety, and time management Behavior by computing the average score for each scale and conducting an Independent Sample T-test to explore possible differences based on gender and degree level.
3. Finally, we developed five structural models, using the Partial Least Squares - Structural Equation Modeling (PLS-SEM) [41], to investigate the relationships between stress, pandemic anxiety, and time management factors, (see Figure 3). This approach is recommended for modelling relationships **between variables** [21].

**Fig. 3.**
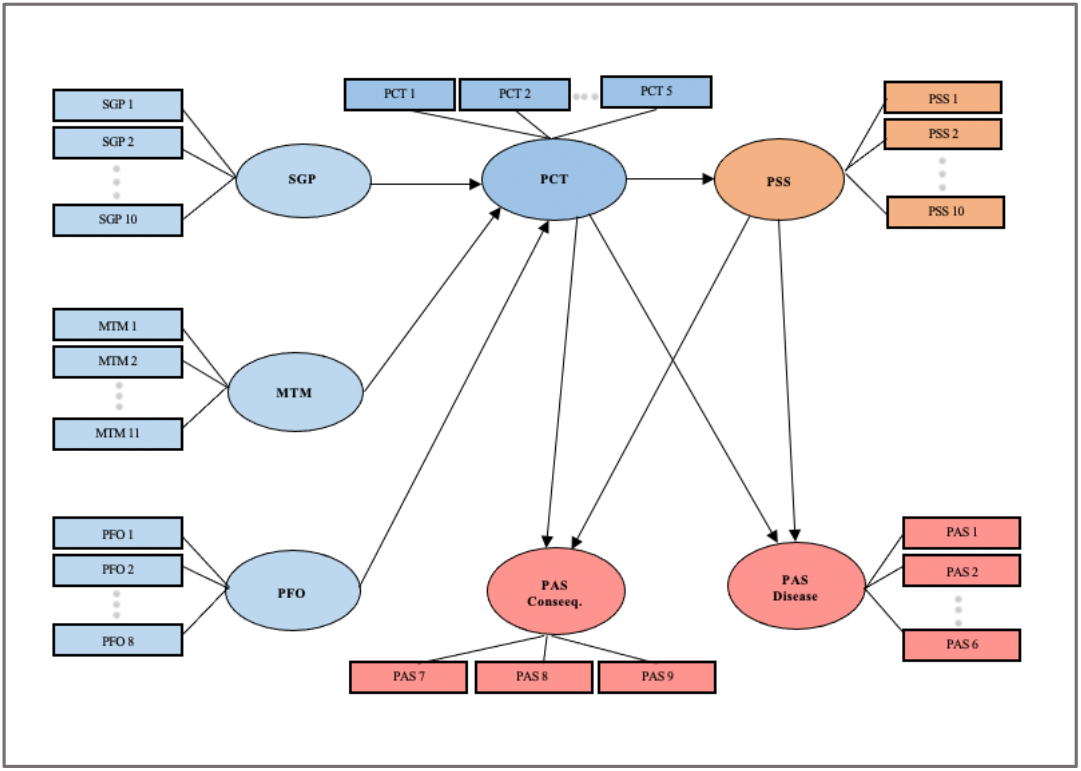
PLS SEM model structure.

### 2.4 Assessing the suitability of data

To determine the suitability of our data, we used KMO sampling adequacies and the Bartlett Test of Sphericity. The result of KMO was 0.891, which is above the recommended value (0.6). Bartlett Test of Sphericity was statistically significant (χ2(1485) = 10373.781, p < 0.0001). These results indicated that our data were suitable for further analysis [21].

### 2.5 Measurement model

Using PLS-SEM, we measured the reliability and validity of a total of 5 models: an overall model for all students and four sub-models (2 gender-based and 2 degree-based models) using the following set of criteria.

- We performed a component-based confirmatory factor analysis (CFA). Each question/indicator was loaded with their corresponding factor. In all models, indicators that had factor loadings of at least 0.5 were kept, and indicators with factor loadings less than 0.5 were removed [10].
- To examine how every indicator strongly correlates with its variables, we measured the reliability of the models using the Cronbach’s alpha and composite reliability. In each model, the Cronbach’s alpha and composite reliability scores were higher than a threshold of 0.7 [10].
- The validity was measured using both convergent and discriminate validity. We used average variance extracted (AVE) to check the data for convergent validity. The results revealed that all constructs AVE scores were higher than the recommended threshold of 0.5 [10]. Heterotrait-Monotrait (HTMT) ratio of correlations was used to measure discriminant validity. HTMT scores for all models were below the recommended limit of 0.9 [10].

## 3 RESULTS

After excluding incomplete and incorrect responses to the survey’s comprehension and attention-determining questions, a total of 502 responses were analyzed. Table 1 presents the results of participants’ demographics. The study sample was diverse in terms of age, gender, current degree of study and country of residence. The majority of participants were undergraduate males from the United States.

**Table 1.**
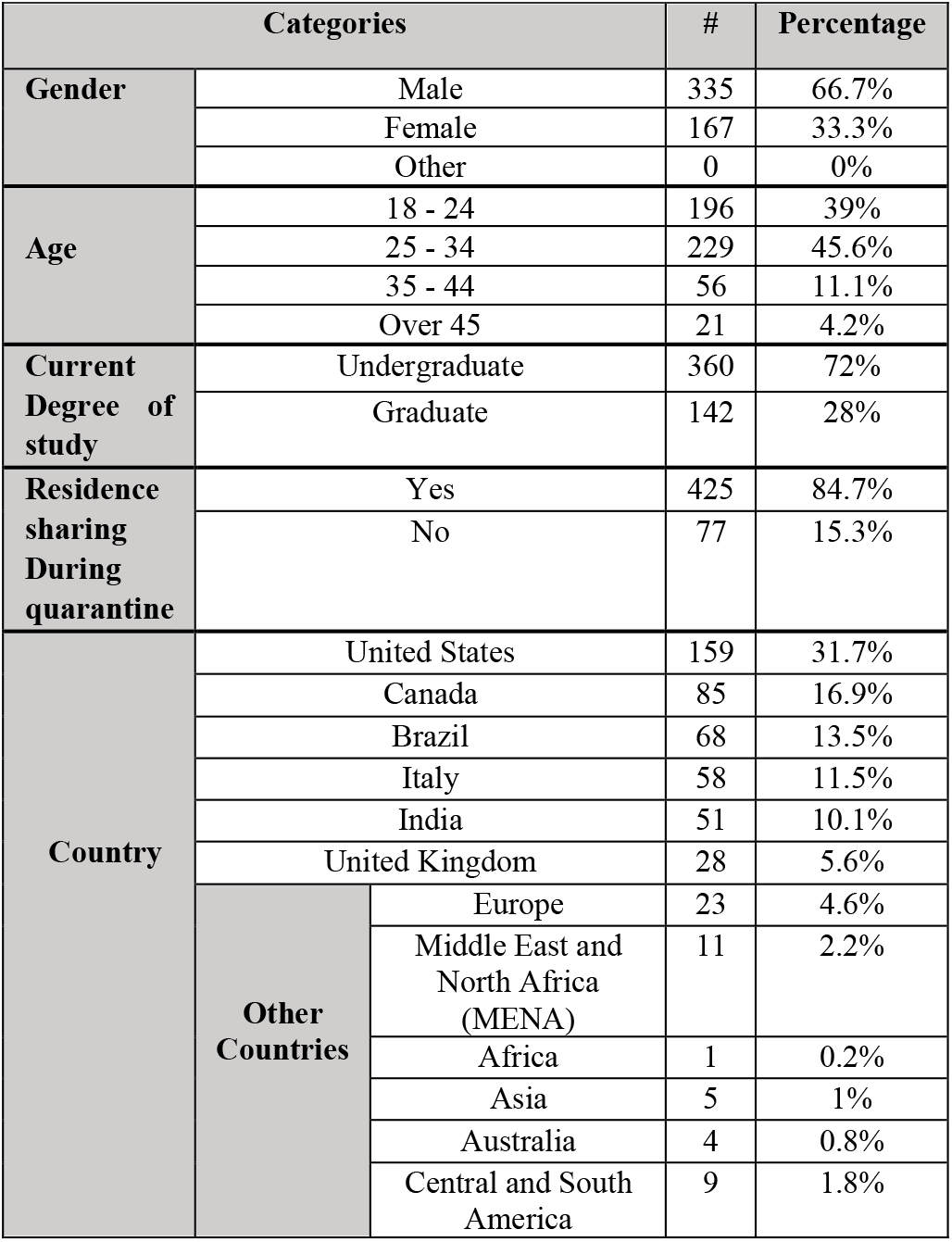
Demographics of the participants

### 3.1 Perceived Stress Scale (PSS-10)

To answer our **RQ1** (*Did students in higher education suffer stress?)* and **RQ2** *(Are there any differences between students’ stress levels based on their degree level or gender?)*, this section presents the results of PSS-10; students’ overall stress levels are presented, then the results will be segregated by gender and degree level.

#### 3.1.1 Overall students’ stress levels and differences between genders

The total average score of stress among students was 20.92 ± 5.14, which is within moderate stress levels. Stress levels by gender are summarised in Table 2. Stress was associated with gender (t(500)= -5.09, p < .0001) as female students experienced more stress than males. Only 9% of male students perceived high stress, while 19% of the total number of females perceived stress in high levels.

**Table 2.** Stress levels by Gender (n = 502)

| Stress Level | Overall<br>n = 502 | Male<br>n = 335 | Female<br>n=167 |
| --- | --- | --- | --- |
|  | Freq. | Freq. | Freq. |
| High | 12% | 9% | **19%** |
| Moderate | **80%** | 82% | 76% |
| Low | 8% | 9% | 5% |

#### 3.1.2 Students’ stress levels by gender and degree level

Students were further divided based on their degree levels. Table 3 shows that 23% of the undergraduate female students reported high stress compared to 8% of the total undergraduate male students (t(358)= -4.69, p < .0001). Only 3% of graduate females in the study reported low perceived stress levels compared to 14% of the total male students in the same degree level (t(140)= -2.17, p < .0001). We further compared females enrolled in undergraduate degrees and those enrolled in graduate degrees; no significant difference was detected.

**Table 3.** Stress levels by degree and gender (n = 502)

| Stress Level | Undergraduate |  |  | Graduate |  |  |
| --- | --- | --- | --- | --- | --- | --- |
|  | Overall<br>n= 360 | Male<br>n = 251 | Female<br>n = 109 | Overall<br>n= 142 | Male<br>n = 84 | Female<br>n = 58 |
|  | Freq. | Freq. | Freq. | Freq. | Freq. | Freq. |
| High | 13% | 8% | **23%** | 11% | 10% | 12% |
| Moderate | 80% | 84% | 72% | 80% | 76% | 84% |
| Low | 7% | 8% | 6% | 10% | 14% | **3%** |

### 3.2 Pandemic Anxiety Scale (PAS-9)

To address the anxiety component of **RQ1** (*Did students in higher education suffer anxiety?*) and **RQ2** (*Are there any differences between students’ anxiety levels based on their degree level or gender?*), this section presents the results of PAS-9. The specific results of the two types of pandemic anxiety (PAS disease and PAS consequences) will also be separated by gender and degree.

#### 3.2.1 Overall results for pandemic anxiety disease and consequences

The overall average of disease-related anxiety among students (PAS disease) was 3.56 ± .75, while the overall average of anxiety driven by consequences (PAS consequences) was 3.64 ± .81. This means that all students suffered from the two types of anxiety during the pandemic. Figure 4 shows the average and distribution results of each type.

**Fig. 4.**
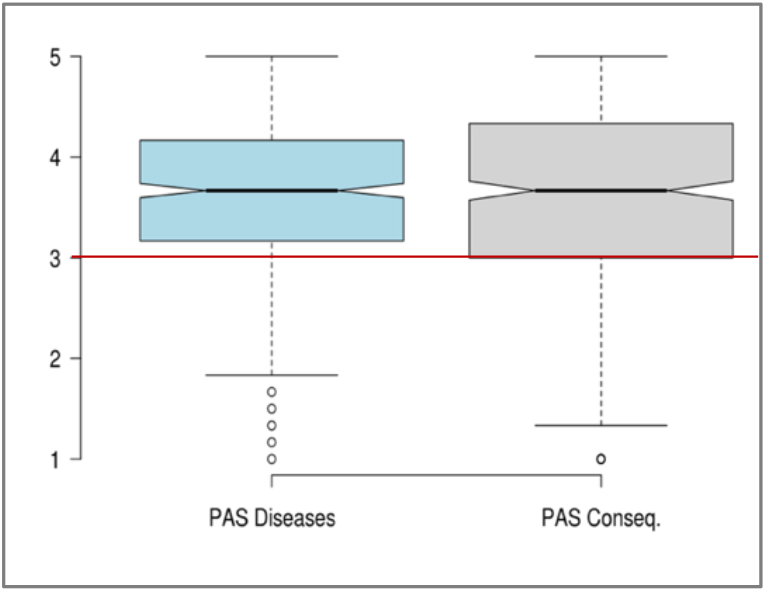
Boxplot of students’ responses (on a scale from 1 to 5, y-axis) to types of pandemic anxiety (PAS-9). Neutral rating is represented by the red horizontal line. Higher rating indicates higher anxiety.

#### 3.2.2 Students’ Anxiety scores by gender and degree level

Table 4 summarizes the results of PAS-9. Students’ results are presented by gender and degree level. Significant relationship was also found between anxiety and gender. Generally, female students reported higher average scores in both types of anxiety, PAS disease (t(500) = -3.26, p < .01) and PAS consequences t(500)= -3.81, (p < .0001), compared to male students. While both undergraduate and graduate students reported similar average scores in PAS disease, graduate students reported a higher average score in the PAS consequences (t(500) = -2.29, p < .05) compared to undergraduate students. Therefore, there is an association between anxiety related to pandemic consequences and students’ degree level. Graduates suffer from higher anxiety as a result of pandemic consequences compared to undergraduates.

**Table 4.** Anxiety level by gender and degree (n = 502)

| PA Types | Mean ± SD | | | |
| --- | --- | --- | --- | --- |
|  | Gender<br>n = 502 |  | Degree level<br>n = 502 |  |
|  | Male<br>n = 335 | Female<br>n = 167 | Under-graduate<br>n = 360 | Graduate<br>n = 142 |
| Disease | 3.50 ± .77 | **3.69 ± .70** | 3.56 ± .76 | 3.58 ± .83 |
| Conseq. | 3.54 ± .82 | **3.83 ± .75** | 3.57 ± .73 | **3.77 ± .75** |

### 3.3 Time Management Behavior (TMB) factors

To address **RQ3** (*What are the most practiced time management Behaviors among students?*), Figure 5 shows the results of the four factors of the TMB scale. SGP was the most practiced factor among students (*M*=3.54 ±.69); this was followed by MTM, which included making lists, planning, and scheduling (*M*= 3.09 ±.78), and then by PFO factor (*M*=3.03 ±.88). SGP and MTM are considered behavioral factors. PFO factor contains a mix of behavioral and perceptual items that are intended to measure students’ preference for organizing their tasks and study spaces. The least expressed factor was PCT (*M*= 2.83 ±.87); this is a perceptual factor, which indicates that students had a low perceived ability to control time during the pandemic.

**Fig. 5.**
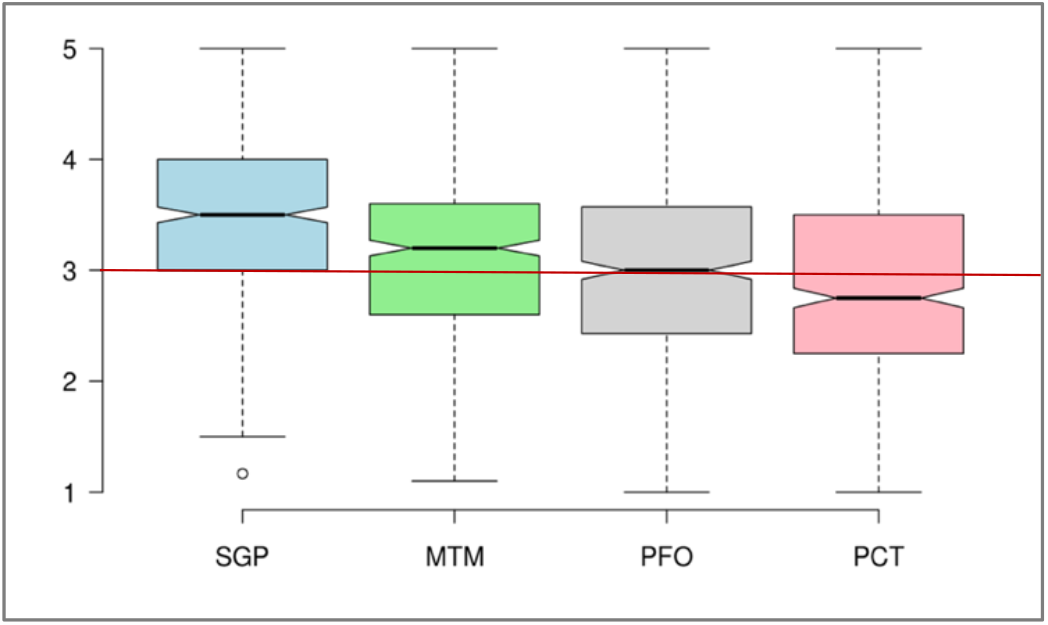
Boxplot of students’ responses (on a scale from 1 to 5, y-axis) towards each of the four TMB factors (x-axis). Neutral rating is represented by the red horizontal line. Higher rating indicates higher practice. **SGP:** Setting Goals/Priorities. **MTM:** Mechanics of Time Management. **PFO:** Preference for Organization. **PCT:** Perceived Control of Time

### 3.4 Relationships between TMB, Perceived Stress, and Pandemic Anxiety during COVID-19 Pandemic

To address **RQ4** (*Which time management factor is significantly associated with perceived control over time*?) and **RQ 5 (***Are there any gender- and degree-level differences in the relationships between stress, anxiety, and various time management factors*?), we developed structural models using PLS-SEM. The level of path coefficient (β) and the significance of path coefficient (p) were used to assess the strength of the relationships between variables [38].

Figure 6 summarizes the path coefficients (β) and their corresponding significance levels (p) obtained from the overall structural model. The model revealed significant positive relationships between PFO and PCT (β = .65, p<.001). A negative relationship was found between PCT and PSS (β= -.33, p<.001). Furthermore, we found a positive relationship between PSS and both types of pandemic anxiety, PAS disease and PAS consequence, (β= .38, p<.001) and (β= .37, p<.001), respectively.

**Fig. 6.**
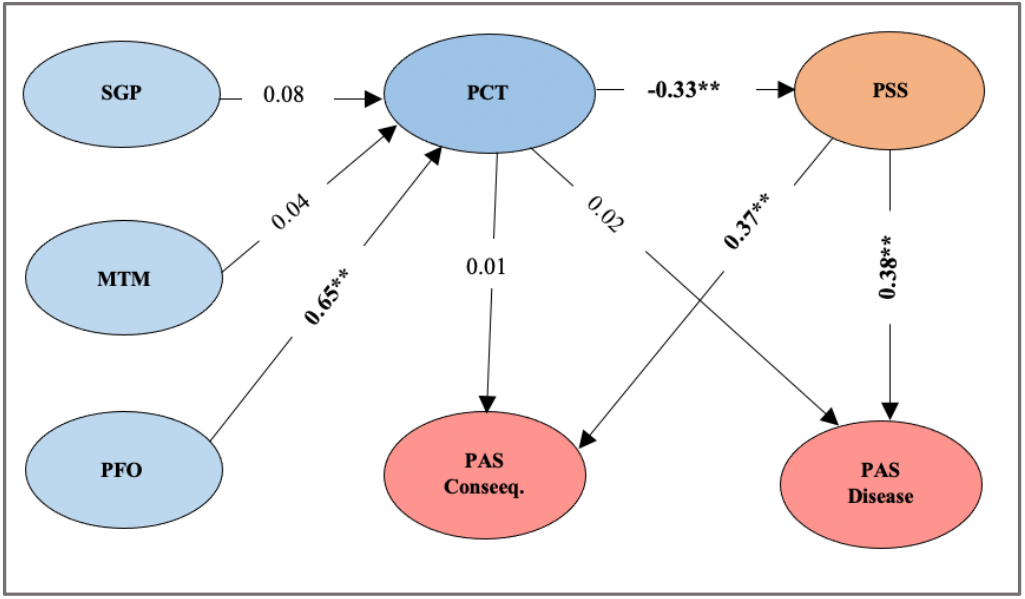
Standardized path coefficients and significance of relationships in the overall model. \*\***Bolded** coefficients are p<.0001, non-Bolded are non-significant. **SGP**: Setting Goals/Priorities. **MTM**: Mechanics of Time Management. **PFO**: Preference for Organization. **PCT**: Perceived Control of Time. **PSS**: Perceived Stress Scale. **PAS**: Pandemic Anxiety Scale.

### 3.5 Moderating effect of gender and degree level between stress, anxiety, and TMB factors

The path coefficients (β) and their corresponding levels of significance (p) were obtained from the gender-based structural model. The model revealed significant results similar to those of the overall model. Significant positive relationships between PFO and PCT were found in both female (β = .58, p<.0001) and male (β = .71, p<.0001) models. Significant negative relationships were found in both gender-based models between PCT and PSS, for female (β= -.31, p<.0001) and for male (β= -.35, p<.0001). Moreover, we found positive relationships between PSS and PAS disease in the female and male models, (β= .34, p<.0001) and (β= .38, p<.0001), respectively. Positive relationships were also found between PSS and PAS consequences for both genders. See Figure 7.

**Fig. 7.**
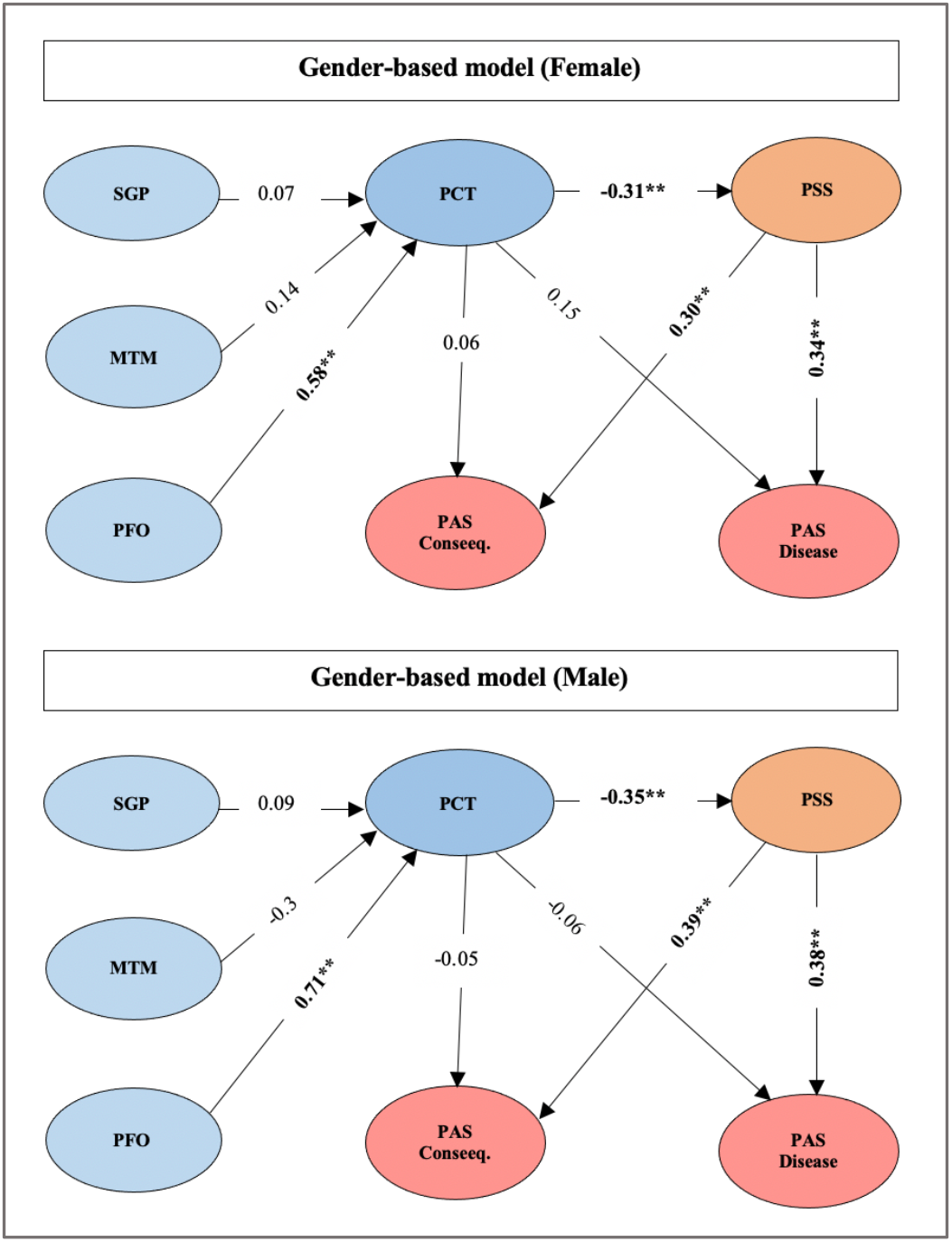
Standardized path coefficients and significance of relationships in the Gender-based model. \*\***Bolded** coefficients are p<.0001, non-Bolded are non-significant. **SGP**: Setting Goals/Priorities. **MTM**: Mechanics of Time Management. **PFO**: Preference for Organization. **PCT**: Perceived Control of Time. **PSS**: Perceived Stress Scale. **PAS**: Pandemic Anxiety Scale.

Figure 8 presents the results of the degree-based model. The model revealed significant results similar to those of the gender and overall models. Significant positive relationships between PFO and PCT were found in both undergraduate and graduate levels (β = .54, p<.001) (β = .82, p<.0001). Significant negative relationships in both degree levels were found between PCT and PSS, in the undergraduate (β= -.29, p<.0001) and graduate (β= -.41, p<.0001) models. Furthermore, we found positive relationships between PSS and PAS disease in the undergraduate and graduate models (β= .41, p<.0001) and (β= .32, p<.001), respectively. Positive relationships between PSS and PAS consequences were found in both undergraduate (β= .42, p<.0001) and graduate (β= .30, p<.0001) models.

**Fig. 8.**
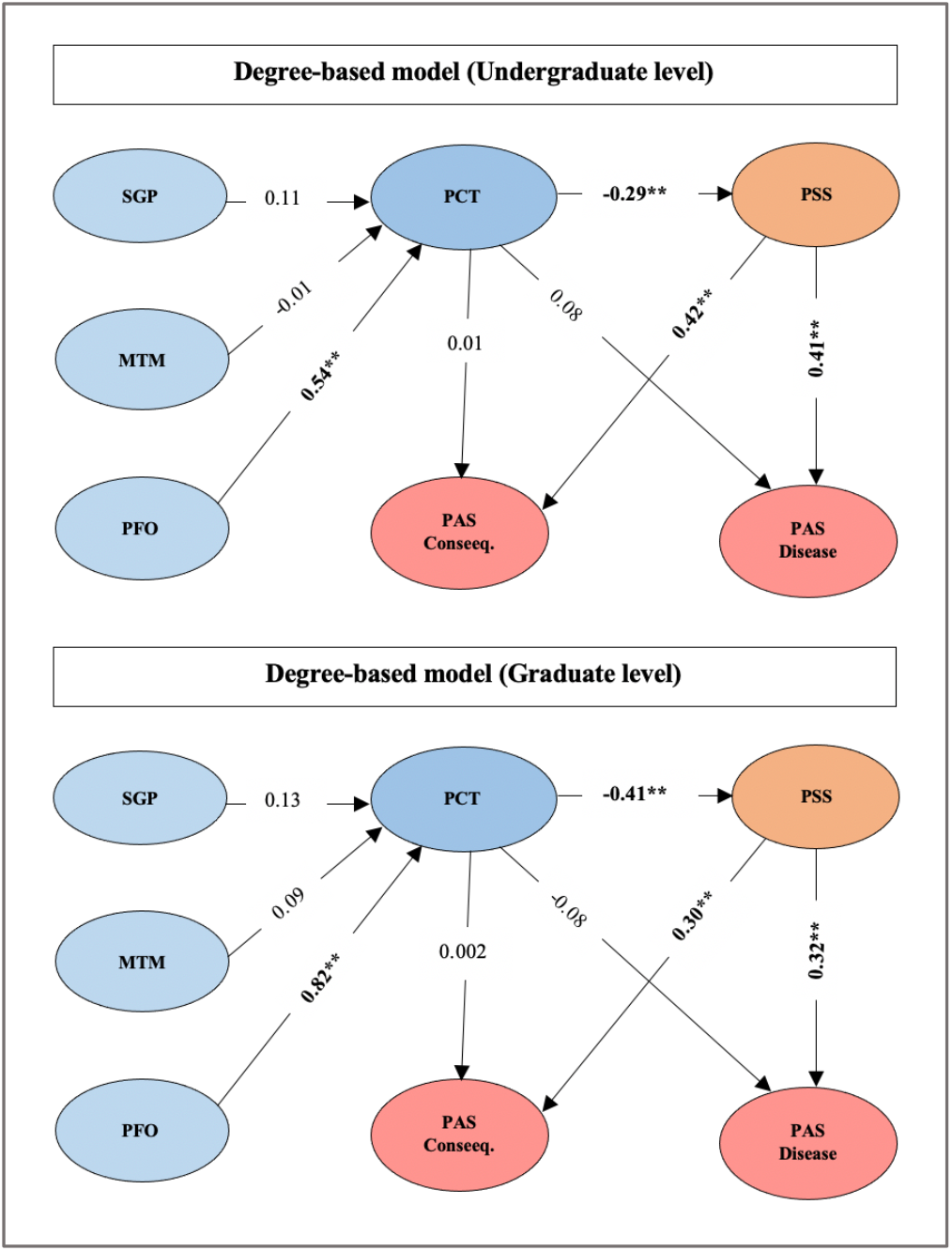
Standardized path coefficients and significance of relationships in the Degree-based model. \*\***Bolded** coefficients are p<.0001, non-Bolded are non-significant. **SGP**: Setting Goals/Priorities. **MTM**: Mechanics of Time Management. **PFO**: Preference for Organization. **PCT**: Perceived Control of Time. **PSS**: Perceived Stress Scale. PAS: Pandemic Anxiety Scale.

## 4 DISCUSSION

Students in higher education make up a population that is susceptible to mental health issues. Our study focusses on the mental health and time management Behavior of students in higher institutions during the COVID-19 pandemic. The findings show that during the pandemic, students suffered from moderate stress and two types of pandemic-related anxiety: anxiety driven by worries about the disease itself and anxiety related to other consequences of the pandemic.

Generally, female students had higher stress levels compared to male students. Although “fight-or-flight” is a common characterization of human physiological and behavioral response to stress [1][47], females are more likely to respond differently. With a behavioral response that manifests in a pattern of “tend and befriend”, females tend to create and maintain a social network that may aid in this process [50]. During the pandemic, the life-threatening situation and the implementation of strict public health measures, including the requirement of social distancing and self-quarantine, have affected social relationships between people and their empathy towards others [44]. In such situations, social support has become limited, which could have contributed to higher stress among female students [50]. Compared to male students, female students also reported higher averages in both types of anxiety (disease and consequences). Stress is a common trigger for anxiety [48], which explains the difference in anxiety levels between genders. Another possible explanation of why females report higher stress and anxiety is gender inequality in social roles, which allocates most household chores to females [8]. Females are primarily responsible for caring for children, ageing parents, and family members [14]. When everyone stayed at home during the lockdown, these chores combined with regular study demands could have increased female students’ stress and anxiety.

Furthermore, our results show that graduate students were more anxious about the consequences of the pandemic than undergraduate students. A possible reason is that graduate students are generally more mature and responsible, many of whom may have additional obligations towards their families [13]. Graduate students who are often more matured shoulder more responsibilities compared to undergraduate students. Some of them may be married with family and hence more anxious about the possible consequences of the pandemic not only on themselves but on their immediate family members (kids and husbands). Because they are also matured, they may also be worried about the possible impact on the job market and their ability to complete their studies and secure a good job upon graduation to take care of themselves and other responsibilities compared to undergraduate students. The study findings indicate that they were more anxious about the finances and the long-term impact of the pandemic on their job prospects [13].

In addition, most graduate students who participated in the research were greatly impacted. A recent study reported that more than 71% of graduate students did not participate in courses during the pandemic; they simply could not do as much due to the decision of pausing experiments and closing down research laboratories [49]. On the other hand, undergraduate students were less anxious as they are generally younger and often have no dependents and as much responsibility outside school.

Regarding the time management behavior factors as defined by Macan [26], students reported *goals setting* and *task prioritization* as common strategies (SGP) used. The practice of other time management strategies such as *planning* and *scheduling* was also reported (MTM). However, the majority of students perceived low ability to control their time (PCT). The low perceived control over time may be attributed to the sudden change in students’ lifestyles during the pandemic, including the shift in their education from face-to-face to online. This disruption was accompanied by an urgent need to adjust by making substantial changes to students’ daily schedules (and behavior). A recent report on the impact of online learning during the pandemic attributed students’ mental health concerns to the isolation from other students and instructors, lack of guidance and counseling, and the difficulty and distractions associated with the use of online platforms [2]. Such considerable changes can explain why it appears that they were not in control of their time.

### 4.1 Relationships between stress, anxiety and time management Behavior factors

Nonis et al. [32] strongly correlated lower academic stress levels with high perceived control over time. Our findings prove that students’ perceived control over time has a legitimate ability to control general stress even during COVID-19 critical times. Since stress and anxiety go hand in hand, improved perceived control over time is pivotal to reducing both.

Prior to the pandemic, studies investigating time management factors, found that setting goals and priorities (SGP) and preference for organization (PFO) were significant in promoting perceived control over time [27]. In this study, all five models, which were generated to examine the differences between students based on genders and degree levels, revealed similar results; students’ preference for organization (PFO) was the only significant factor that influenced students’ perception of the ability to control time. The sudden closure of universities, campus libraries, and shared study spaces has placed students in a chaotic situation, yet, in complete charge of their time and learning.

Lacking an established approach to task and study space organization (preference for organization, PFO), is the factor that strongly influences the perception of controlling time. Students who are able to manage time through the organization of tasks and study spaces had a higher perception of control over their times, hence, less stress and anxiety. It has been reported that physical clutter in study space leads to mental clutter [34]. McMains and Kastner [30] indicated that disorganization and clutter can increase cognitive overload and reduce the ability to focus. Another research reported that higher cortisol (stress hormone) levels were found among individuals in cluttered environments [45]. All these explains why organization of task and space is strongly associated with perceived control of time and hence reduce and anxiety.

### 4.2 Toward persuasive intervention design

Organization is one of the keys to success in accomplishing goals. Being organized can help students have a clear picture of what is expected to be completed and when. The preference for organization factor (PFO) is comprised of six perceptual and behavioral items, which can be grouped into three main categories: *1. items related to the organization of tasks, 2. items related to the organization of space, and 3. items related to the benefits of being organized*. These categories encompass the concept of organization in students’ daily lives.

Organization is essential, especially when students are fully responsible for their educational process and environment. Therefore, after examining the literature, brainstorming, and agreement between experienced researchers specialized in the field of persuasive technology, we have mapped the PFO categories to actionable persuasive strategies, which can be used in designing persuasive interventions to assist students’ and promote positive perceptions and attitudes towards task and space organization. The mapping was based on the Goal-Setting strategy [25] and the Persuasive Systems Design (PSD) framework [33]. (See section 1.2).

**Table 9.** Mapping of PFO factor to corresponding persuasive strategies based on the PSD framework and goal-setting strategy.

| PFO categories | Persuasive Strategies |
| --- | --- |
| Task organization | *Goal setting, reduction, self-mentor* |
| Space organization | *Suggestion* |
| Benefits of being organized | *Rewards, social learning, reminder* |

### 4.2.1 Task organization

This category of task organization involves actions such as scheduling the day, having to do list, and starting with important tasks first. Several persuasive strategies can be used to help students organize their tasks. Task organization is not easy to develop and maintain. The use of ***goal-setting strategy*** would help students direct their attentions and increase their task performance. Making goals incrementable may also help them build confidence in their performance [37].

The use of ***reduction strategy***, through which a complex Behavior is deconstructed to simple tasks to aid users in performing the target Behavior [33]; is important to minimize students’ efforts in setting and organizing their goals. The system can provide some templates for organizing and scheduling tasks which students can easily fill or change as necessary. Since feeling overwhelmed can affect deciding what is important and urgent and what is not, the system should automatically arrange and present students’ goals according to their importance/urgency.

The use of ***self-mentoring strategy*** allows students to observe and track their performance and progress toward their specific goals. Self-monitoring enables students to distinguish between realistic and unrealistic goals and encourages them to identify and foster productive work habits and better control of their time.

#### 4.2.2 Space organization

This category includes actions such as cleaning and organizing the study space, for example, at the end of the daily study. Informing and encouraging students to adopt such organizational approaches can be achieved using a ***suggestion strategy***. Offering fitting suggestions has been reported to have greater persuasive power [33]; the system should therefore provide suggestions and tips to help students minimize physical clutter to create a better-organized study space.

#### 4.2.3 Benefits of being organized

This category speaks to the benefits of being organized. Students should be aware of the value of being organized. Previous studies suggest that positive reinforcement and gain-framed appeal have the potential to operationalize perceived benefits in persuasive technology design [37] [51] [52]. Positive reinforcement [52] can be accomplished by rewarding completed tasks and goals (***rewards strategy***) using virtual rewards such as badges or points that can be collected toward redeemable values. Gain-framed messages that emphasize the benefits of adhering to organizational acts [51] can be implemented using ***reminders/notifications strategy***. An example of gain-framed messages is *“When you are organized, you’ll be able to better adjust to unexpected events”*.

Moreover, ***social learning strategy*** can increase users’ motivation toward the target Behavior by observing the outcomes of others who are performing the same Behavior [33]. The use of this strategy would allow students to share insights into effective organizational strategies. The application of this strategy is possible through incorporating a shared journal in the system design. The shared journal would provide opportunities for students to observe, interact, and learn new organizational strategies and techniques posted by other students.

## 5 CONCLUSIONS

The study objective was to explore the impact of the COVID-19 pandemic on students’ mental health and time management behavior. Specifically, we examined the relationships between stress, anxiety, and different time management factors during the COVID-19 pandemic. Results from a large-scale study of 502 participants show that students suffered from both stress and anxiety. Female students appeared to be more vulnerable to stress and both types of pandemic related anxiety. Graduate students were more anxious about the pandemic consequences than undergraduate students. Students’ preference for organization (PFO) was the only factor that significantly influenced their perceived ability to have control over time (PCT), which contributes to reducing stress, hence, anxiety. To promote students’ preference for organization, we map the three categories of organization to corresponding persuasive strategies which could be used in the design of persuasive interventions. This creates an opportunity for developing interventions to improve students’ perceptions of control over time, thus, less stress and anxiety.

## Data Availability

The dataset of this manuscript/study is not available online as it is restricted by an ethics application issued from Dalhousie University.

## 6 ACKNOWLEDGEMENT

This research was undertaken, in part, thanks to funding from the Canada Research Chairs Program. We acknowledge the support of the Natural Sciences and Engineering Research Council of Canada (NSERC) through the Discovery Grant.

